# NASCarD (Nanopore Adaptive Sampling with Carrier DNA): A rapid, PCR-free method for whole genome sequencing of pathogens in clinical samples

**DOI:** 10.1101/2023.03.10.23287094

**Authors:** Miguel A. Terrazos Miani, Loïc Borcard, Sonja Gempeler, Christian Baumann, Pascal Bittel, Stephen L Leib, Stefan Neuenschwander, Alban Ramette

**Affiliations:** Institute for Infectious Diseases, University of Bern, Bern, Switzerland

**Keywords:** Beta coronaviruses, SARS-CoV-2, whole-genome sequencing, next generation sequencing, genomics, nanopore

## Abstract

Whole-genome sequencing (WGS) represents the main technology for SARS-CoV-2 lineage characterization in diagnostic laboratories worldwide. The rapid, near-full-length sequencing of the viral genome is commonly enabled by high-throughput sequencing of PCR amplicons derived from cDNA molecules. Here, we present a new approach, called NASCarD (Nanopore adaptive sampling with carrier DNA), which allows low amount of nucleic acids to be sequenced while selectively enriching for sequences of interest, hence limiting the production of non-target sequences. Using clinical samples positive for SARS-CoV-2 during the Omicron wave, we demonstrate how the method leads to up to >100x coverage of the full genome sequences of the target organism as compared to standard shotgun metatranscriptomics approach. It provides complete and accurate genome sequence reconstruction within seven hours at a competitive cost. The new approach may have applications beyond SARS-CoV-2 sequencing for other DNA or RNA pathogens in clinical samples.

## INTRODUCTION

Severe Acute Respiratory Syndrome Coronavirus 2 (SARS-CoV-2) is a positive-strand RNA virus of the Betacoronavirus genus from the Coronaviridae family, which was discovered in the region of Wuhan in December 2019 (Zhu et al. 2020). Due to the rapid spread of the COVID-19 associated diseases, a worldwide effort has been conducted to monitor the emergence and evolution of SARS-CoV-2 lineages due to the implications these new variants may have for public health, society, and scientific research (Somerville et al. 2021). While the detection of variants of concern in clinical samples could be done using polymerase chain reaction (PCR; e.g. (Vogels et al. 2021; Bedotto et al. 2021)), most laboratories have adopted Next Generation sequencing (NGS) approaches, using either short-read and long-read sequencing methods (Liu et al. 2021; Freed et al. 2020), to fully characterize the genomic information of circulating lineages. Among these methods, the ARTIC protocol was developed based on an earlier strategy for sequencing single-stranded RNA viruses from high cycle threshold (Ct) clinical samples (Quick et al. 2017). The protocol consists of PCR-tiling of short (∼400-bp) or long (2-2.5-kb) amplicons followed by NGS of the amplicons (Plitnick et al. 2021). The consensus genomic sequences are then taxonomically classified using NextClade (Hadfield et al. 2018) or the dynamic approach of Pangolin (Rambaut et al. 2020). Amplicon-based sequencing is among the most popular methods as it provides an efficient and cost-effective solution to massive sequencing needs while allowing the sequencing at low concentrations of target cDNA (Charre et al. 2020; Vrancken et al. 2016). Mutations at primer binding sites may yet impact the ability to generate near-complete genomes when using amplicon-based sequencing approaches, leading to amplicon drop-offs, e.g. (Kuchinski et al. 2022; Borcard et al. 2022). Thus, PCR primer sequences and concentrations must be frequently updated to ensure the correct characterization of new circulating variants, which can represent a challenging task, e.g. (Davis et al. 2021; Lambisia et al. 2022). Besides PCR-specific approaches, random priming amplification combined with WGS has been shown to successfully recover RNA viral genetic material from SARS-CoV-2 (Chrzastek et al. 2022). Yet, the method may also be challenged by the presence of non-specific bacterial or host genes in clinical samples, which can greatly reduce the efficiency of viral genome amplification and sequencing, and still require the optimization of the PCR condition to improve target recovery (Chrzastek et al. 2017).

Shotgun metagenomics is a PCR-free approach, which does not require prior knowledge of the target genomic sequence. Applications in routine diagnostic laboratories may still be hampered by its costs, availability of sequencers, and by its lack of sensitivity when multiple species, including host DNA or RNA, may be present (Marotz et al. 2018). Others have used direct RNA sequencing (DRS; Oxford Nanopore Technologies, ONT) to sequence corona viruses natively (Viehweger et al. 2019). While DRS may be readily applied as a shotgun metatranscriptomic approach to the total RNA extract from a clinical sample e.g. (Grädel et al. 2020), it also has specific requirements: Because only polyadenylated RNA molecules are sequenced, a high amount of RNA material is necessary for ONT library preparation and is still costly due to the current lack of sample multiplexing possibilities. This approach is thus not suited for routine clinical work with SARS-CoV-2 positive samples (Vacca et al. 2022). Therefore, there is an urgent need to develop new approaches to obtain unbiased high-quality sequences of SARS-CoV-2 at a competitive cost.

ONT sequencers, which offer many advantages for sequencing long DNA molecules in real-time from complex samples at competitive prices (Y. Wang et al. 2021), including clinical samples (Petersen et al. 2019; Neuenschwander et al. 2020), have recently enabled the targeted sequencing of specific DNA or cDNA molecules, a process termed “Adaptive Sampling” (AS; previously known as “Read Until”; (Loose, Malla, and Stout 2016)). In this application, a binary decision step can be implemented at the pore level during sequencing, allowing either the sequencing to completion or the rejection of a DNA molecule, by reversing the driving voltage at the nanopore level (Oxford_Nanopore_Technologies 2020; Kovaka et al. 2020; Payne et al. 2021). This method is proving to be effective for identifying pathogens in metagenomic samples, even when the amount of host DNA present is substantial compared to the target pathogen DNA (Kipp et al. 2021; Lin et al. 2022; Martin et al. 2022). However, one recognized limitation of ONT-based library preparation is the need for relatively high amounts of nucleic acid material (Wang et al. 2021; Heavens et al. 2021), as using the ONT 1D ligation technology generally requires a high (typically 1,000 ng or more) amount of genomic DNA (ONT Ligation sequencing gDNA protocol SQK-LSK109; version 25 May 2022).

Our study presents a novel technique called “Nanopore Adaptive Sampling Carrier DNA” (NASCarD) for PCR-free targeted genomic sequencing of organisms of interest. As illustrated here for SARS-CoV-2 genome sequencing in clinical samples, the approach leverages the combined use of i) a genomic carrier to increase the molecular mass of the nanopore input library, thus addressing the potential loss of target cDNA during library preparation, and ii) AS targeted sequencing to enrich for SARS-CoV-2 sequences in clinical samples. Our results demonstrate that this combination provides better and faster genomic coverage than standard nanopore sequencing techniques at a competitive price.

## METHODS

### Sample collection, RNA extraction and cDNA synthesis

Twenty-two viral RNA samples were randomly collected from 5 October 2021 to 18 May 2022 during routine sequencing of SARS-CoV-2 viral genomes performed at the Institute for Infectious Diseases (IFIK, Bern, Switzerland) for diagnostic or surveillance purposes. Ethical approval was granted by the Cantonal Ethical Commission for Research, Canton of Bern, Switzerland (GSI-KEK, BASEC-Nr 2022-00597) to sequence and genomically compare SARS-CoV-2 isolates from samples previously screened and submitted to IFIK for viral diagnosis by treating physicians. Briefly, total nucleic acids were extracted from 200 μl nasopharyngeal swab samples in transport media using the STARMag 96 ×4 Universal Cartridge Kit on a Seegene STARlet liquid handling platform (Seegene, Seoul, South Korea) and eluted in 100 μl of elution buffer according to the manufacturer’s instructions. Cycle threshold (Ct) values for all samples were determined in our facility during routine real-time PCR procedure (cobas SARS-CoV-2 test, Roche, Basel, Switzerland). All test results were anonymized prior to sequencing and further bioinformatic analyses. Nucleic acid eluates were immediately stored at -80°C after processing until further use. Genome sequence information for all isolates is available in the GISAID database (https://www.gisaid.org; Shu and McCauley 2017); and was deposited online (doi: 10.5281/zenodo.7692956). First-strand cDNA synthesis was performed from 16 - 32 μl of nucleic acid extracts using the LunaScript RT SuperMix kit (New England Biolabs, Ipswich, MA, USA). The manufacturer’s protocol was followed by adapting the total reaction volume to the input volume. This step was followed by NEBNext Ultra II non-directional second-strand RNA synthesis (New England Biolabs), and purification using CleanNGS magnetic beads (CleanNA, Waddinxveen, The Netherlands) at a ratio of 1.8x in 40 μl nuclease-free water (New England Biolabs) according to the manufacturer’s protocol.

### Nanopore sequencing

Routine WGS of all RNA extracts presented in the study was done following the rapid protocol for whole-genome sequencing of SARS-CoV-2 on the GridION X5 device. Briefly, this protocol generates 1200-bp tiled amplicons (Freed et al. 2020), which are then further sequenced using a rapid, transposase-based ONT library (SQK-RBK110.96) preparation from Oxford Nanopore Technologies, Oxford, UK (ONT). For the NASCarD approach, library preparation was performed from 40 μl of SARS-CoV-2 ds-cDNA mixed with 8 μl (400 ng) of Enterobacteria phage λ DNA (Lambda DNA) using ONT’s SQK-LSK109 kit following the genomic DNA ligation protocol (version: GDE_9063_v109_revAP_25May2022). In experiments where a control flow cell was run in parallel, a variation of the protocol was performed at the final step, which consisted of using 24 μl of EB (elution buffer) from the kit itself to elute the library in order to obtain a sufficient volume to load the same library preparation onto two different flow cells to be sequenced in parallel. For each sample, prepared libraries were loaded onto two Nanopore R9.4.1 flow cells, one for AS and one for control sequencing, and both flow cells were run at the same time on a GridION X5 device (MinKNOW version 21.05.8) for 6-72 hours.

The output of AS sequencing consists of nanopore reads in a FASTQ format, accompanied by a csv file that lists the classification of each read made by the MinKNOW version of ONT ReadUntil API (https://github.com/nanoporetech/read_until_api) based on read matching to the user-provided reference sequence(s) (**Figure 1**). The whole process consists of the following steps: a) The initial 400-600 bases of a strand that translocates through a given pore (b) are used by the software (c) to classify the read as “enriched” (flagged as “stop receiving” by ONT’s ReadUntil software) if the read matches the reference(s) sequence and is then further sequenced (d). e) A read is rejected (“unblocked”) if its initial bases do not match the reference sequence(s). A voltage inversion at the pore level expels the read out of the pore and prevents its further sequencing. A read may be classified as “no decision” if the read sequence does not match the reference unambiguously. A set of nine SARS-CoV-2 reference sequences were initially chosen to maximize the chances to recover divergent variants and recombinants, which consisted of the original Wuhan-Hu-1 isolate (MN908947.3), and a total of eight sequences from circulating isolates in December 2021 from Switzerland (from three cantons and all Delta (B.1.617.2-like) with GISAID numbers EPI_ISL_1941773, EPI_ISL_2017208, EPI_ISL_7888951), three Indian isolates (from the omicron BA.1.1 lineage, EPI_ISL_7876997, EPI_ISL_7877093, and from BA.1.18 lineage with EPI_ISL_7877202), and two isolates from New York, USA (lineage BA.1 EPI_ISL_7887528, and lineage BA.1.15 with EPI_ISL_7887531). All sequences were deposited at doi:10.5281/zenodo.7692939. Sequencing data were deposited at National Center for Biotechnology Information (NCBI) and can be accessed via BioProject PRJNA930601.

**Figure 1.**
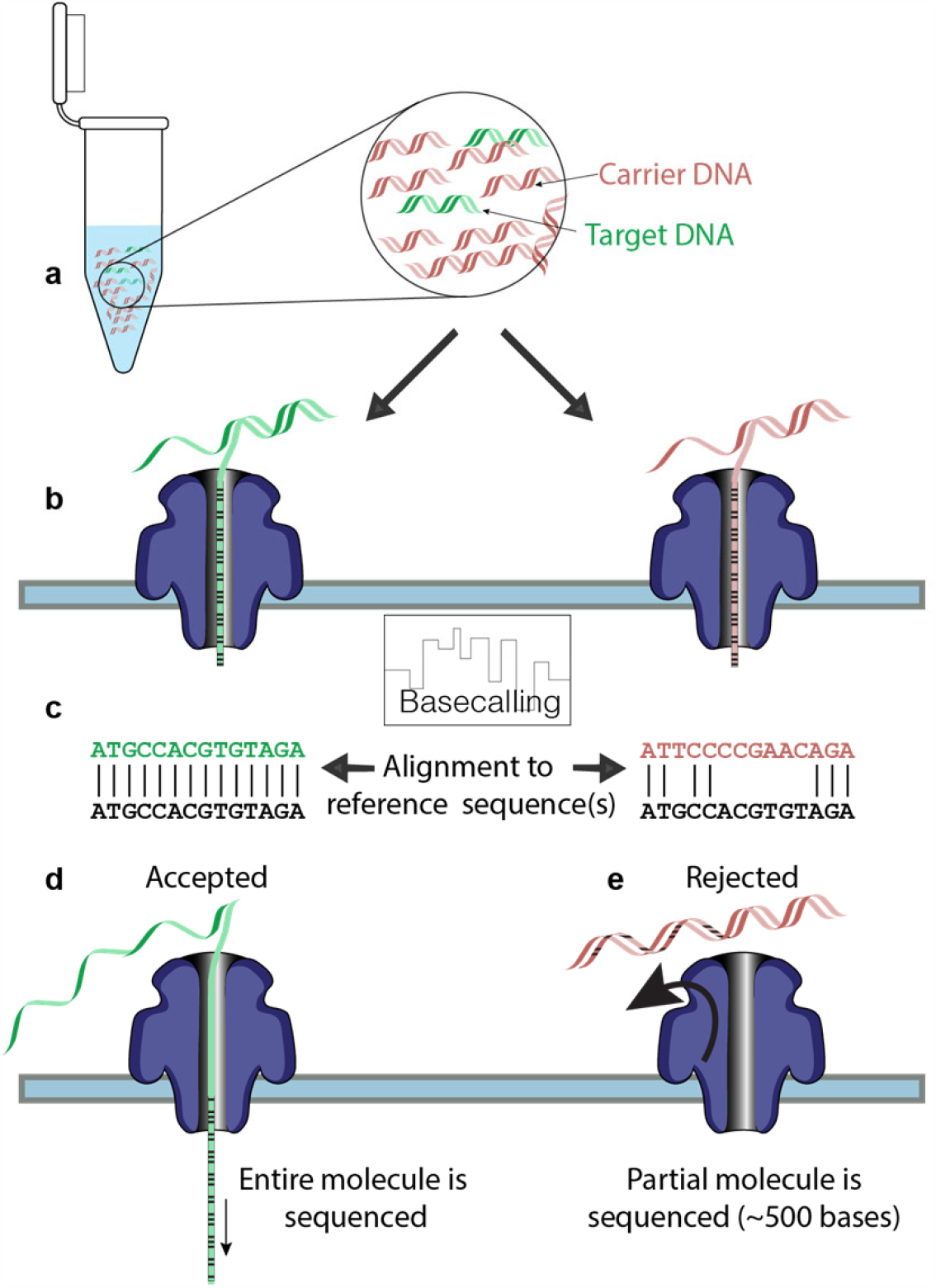
Methodological steps involved in the NASCarD method. a) A mixed sample of target molecules (colored in green) and host molecules (red), is combined with carrier genomic DNA (red) during nanopore library preparation; b) while (c)DNA molecules translocate in the nanopores, and are basecalled into nucleotide sequences, the initial 400-600 bases of the 3’ end of the translocating reads (b) are used by the API to classify the reads based on the read matches to the reference sequence (in black font) (c). Then the decision to further sequence (d) or to reject a read by pore-level voltage inversion (e) is made.

### Bioinformatics Analysis

To confirm the classification made by the ONT software, reads were also independently aligned to the omicron sequence (omicron B.1.1.529; GISAID Accession ID: EPI_ISL_7887531), because it would best match circulating variants at the time of the study. The mapping was performed using *minimap2* (v2.22-r1101). After mapping to the SARS-CoV-2 reference genome, read alignment file was processed further using *samtools* (v1.15; (Danecek et al. 2021)) and unmapped reads to SARS-CoV-2 sequences were further mapped to the Lambda phage and the human genomes (GRCh38/hg38). All basic mapping statistics were further calculated using *seqkit* (v2.2.0, (Shen et al. 2016)) and summarized with the R programming language (version 4.1). All samples from our routine SARS-CoV-2 sequencing (amplicon-based) were routinely analyzed using the ARTIC pipeline (Loman, Rowe, and Rambaut 2020). However, to compare the consensus sequences obtained using the NASCarD method with those from our routine sequencing, we chose to adapt the ARTIC pipeline by removing the primer trimming steps in order to adapt the pipeline to this PCR-free workflow, while keeping all other steps involving variant filtering unchanged (deposited at doi:10.5281/zenodo.7713085). To estimate the flexibility of our approach regarding the emergence of new variants, we opted to create six levels of mutations (5, 10, 15, 20, 30, 50% SNPs) in our reference sequences using *Mutation-Simulator* (Kühl, Stich, and Ries 2021). Nanopore reads produced in experiment V were subsequently mapped to our mutated references using *minimap2* and GNU parallel. The results were analyzed in the RStudio environment.

## RESULTS

### NASCarD improves the completeness and quality of SARS-CoV-2 genome sequences

To evaluate the NASCarD approach, we selected clinical, SARS-CoV-2 positive, RNA samples that were previously successfully sequenced under routine conditions using the ONT rapid WGS 1200-bp amplicon-based protocol. The standard protocol generally provides high-quality sequences (Freed et al. 2020), which we used to evaluate the accuracy and completeness of the newly obtained sequences from our study. We compared the SARS-CoV-2 sequencing performance of NASCarD vs. control sequencing (without AS), in terms of number and average length of SARS-CoV-2 reads in comparison to human and Lambda DNA reads. In total, we performed comparative genomic analyses by considering 23 clinical samples, including two variants (delta, omicron) and recombinant lineages (XM). We evaluated the possibility of multiplexing samples in the same flow cell in two experiments (with 12 and 5 samples, respectively), while, in parallel, performing ten independent sequencing experiments (**Table S1**). From those experiments, we highlight the example of the recombinant SARS-CoV-2 from experiment V (**Figure 2**): After about 18 h of sequencing, a total of 2,989,153 reads were obtained with NASCarD. All reads labeled as “enriched” were confirmed to be on-target by independent mapping to SARS-CoV-2 genome, resulting in a total of 855 reads with an average length of 3,833 bases. The “rejected” reads corresponded almost entirely to either human DNA with 1,324,197 reads or Lambda DNA with 1,128,212 reads. The mean length of human and Lambda reads were 548.2 and 548.7 bases, respectively, as expected for rejected reads (see Methods). Only one “rejected” read with a size of 371 bases was classified as SARS-CoV-2, which was most likely due to the short size of that fragment (**Figure 2A, Table S1**). Meanwhile, the flow cell that ran the same library as a control (i.e. AS was not used) produced a total of 736,061 reads and, of these, only 276 reads mapped to SARS-CoV-2, with an average size of 3,700 bases. This shows that NASCarD can yield approximately up to three times more target reads than a control sequencing run. The average sizes of human and Lambda reads in the control sequencing were 4,561 and 18,216 bases, respectively, thus evidencing the efficiency of NASCarD in preventing the concomitant sequencing of long, non-target molecules (**Figure 2A**). Noticeably, the percentage of sequenced SARS-CoV-2 bases was higher with NASCarD (0.23% of total bases) than in the control (0.01%). The genome coverage of SARS-CoV-2 was expectedly found to be directly related to the number of matching reads (**Figure 2B**). With NASCarD, an average coverage of 109.7x was obtained while in the control it was only of 34.2x (**Figure 2B**). All samples used samples in the study were classified as containing Omicron variants, except one sample that was classified as a Delta variant (“O” experiment; **Table S1**). Similar results as those aforementioned for experiment V (sample V01) were obtained in experiments with either recombinant or non-recombinant SARS-CoV-2 samples, with genome completeness of 99.9% for samples M01 (**Figure S1**), P01 (**Figure S2**), J11 (**Figure S3**), O05 (**Table S1**), and associated Ct from 15.6 to 19.3, obtained within 20 h of NASCarD sequencing (**Table S1**).

**Figure 2.**
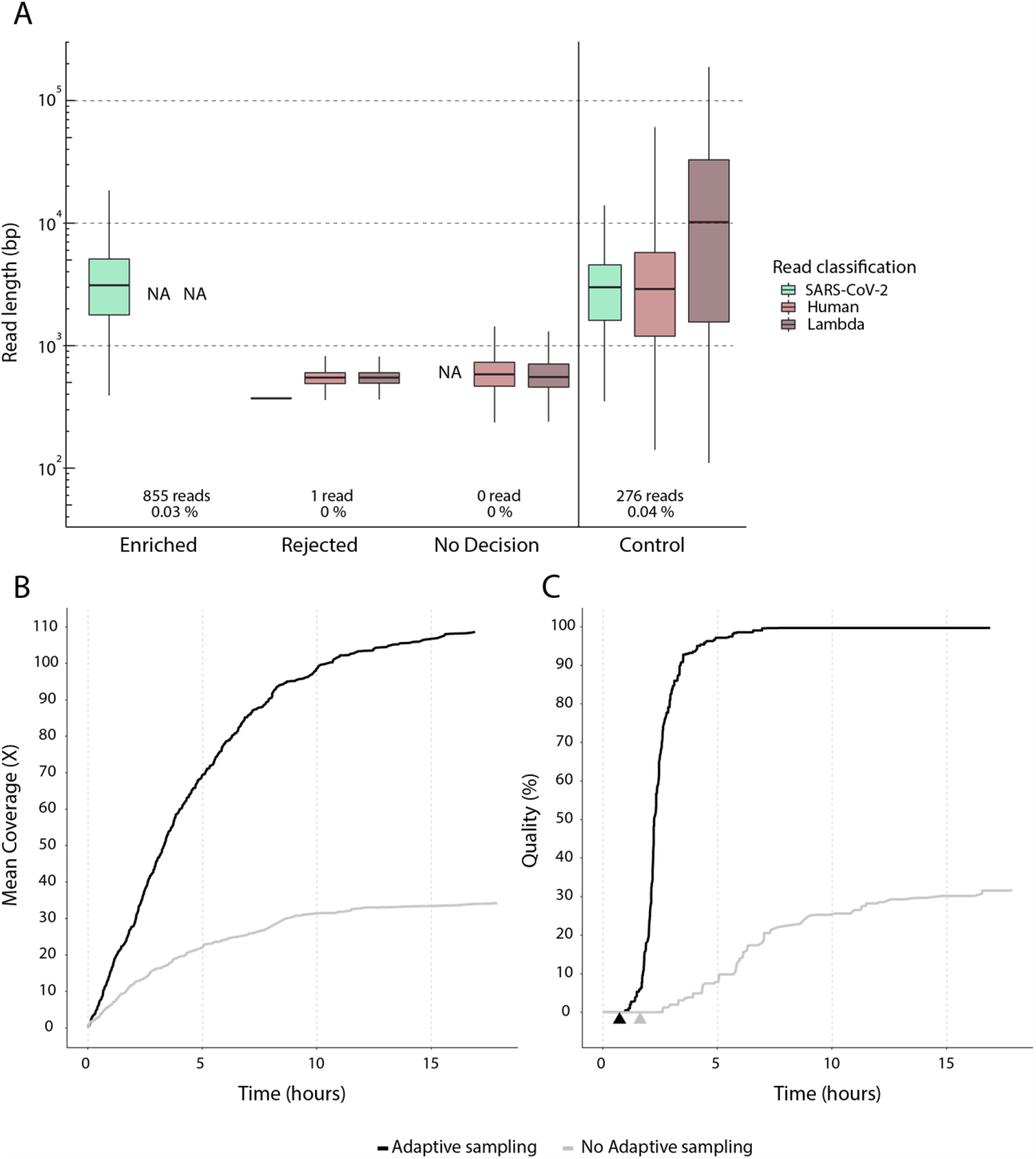
PCR-free, long-read sequencing of SARS-CoV-2 recombinant genome in a clinical sample using NASCarD. A) Read length distribution of the same library preparation for clinical sample V1 with NASCarD and a control run. After each run, all reads were systematically mapped to SARS-CoV-2 genome sequence, Lambda phage and human genomes to validate their correct classification. NA, no read available. B) Comparison of mean coverage for SARS-CoV-2 in AS (black line) and control (grey line) after 18.4 h of sequencing. C) SARS-CoV-2 genome quality (100-N%) over time in AS and control sequencing, where N% represents the percentage of positions with a sequencing depth below 20X. The time when genome completeness reached >99% is indicated for NASCarD (black arrow), and the control flow cell (grey arrow).

### Seven hours suffice to obtain high-quality consensus sequences with NASCarD

To obtain high-quality genome consensus sequences (GCS), we monitored two key parameters: i) Complete recovery of the genome (“completeness”), i.e. the percentage of the total number of covered positions of the original SARS-CoV-2 reference sequence (Wuhan-Hu-1), and ii) sufficient sequencing depth to allow the recovery of a robust GCS (genome quality). We defined “genome quality” as (100-N%), where N% represents the percentage of positions with a sequencing depth below 20x. This definition was based on our observations that 20x sequencing depth is at least needed to recover a good GCS, which is also in accordance with the threshold for minimal sequencing depth required in the ARTIC pipeline (Quick 2020; Quick et al. 2017; Fieldbioinformatics/Artic). With NASCarD, genome completeness reaches >99% in 0.7 h (black arrow), while this took 1.6 h in the control flow cell (grey arrow), i.e., 2.3 times slower (**Figure 2C**). In experiment V, control sequencing produced SARS-CoV-2 GCS quality of only 32% after 18.5 h of sequencing, while AS reached 99.7% in the same timespan (3.15 times higher; **Figure 2C**). Additionally, GCS quality reached >99% after 6.7 h of sequencing using NASCarD, and 99.81% (56 ambiguous or absent bases out of 29,856 bases) after 18.5 h. Comparatively, the quality of the GCS obtained under routine amplicon-based sequencing for the same sample was 99.3% (197/29,770 bases), indicating a superior quality of the GCS produced by NASCarD. When considering the cases when 99.9% completeness was obtained with NASCarD (experiments M, P, V; **Table S1**), NASCarD always produced better quality GCS than the amplicon-based approach: For experiment P, NASCarD yielded 99.87% (37/29,854 bases) vs. 93.94% (1,809/29,873 bases), and for experiment M, again NASCarD produced 99.83% (48/29,858 bases) vs. 96.28% (1,110/29,867 bases), as compared to amplicon-based sequencing.

### GCS quality decreases with lower viral loads

Our results showed that NASCarD produced better GCS quality and higher number of SARS-CoV-2 sequences than control conditions or standard amplicon-based nanopore approaches for Ct <20 (**Table S1**). We next evaluated the performance of AS sequencing on multiple samples with different Ct values, either as multiplexed samples or a single sample per flow cell. SARS-CoV-2 RNA samples with a wide range of Ct values (between 15.5 to 28.3) (**Table S1**) were selected to evaluate the NASCarD approach more generally. Two runs were done with multiplexed samples, one with twelve samples (experiment J) and one with five samples (experiment O). Except for these two multiplexed runs, all samples were processed independently on single flow-cells (experiments M, P, N, V, H, I). The total number of reads mapping to SARS-CoV-2 was 1,296 for the sample with the lowest Ct (15.5), while the sample with the highest Ct (Ct 28.3) produced only four SARS-CoV-2 reads. Since Ct qPCR values may be inversely proportional to the viral load in a sample, we evaluated the relationship between completeness (i.e., the number of bases covered, regardless of coverage) and Ct values (**Figure 3**). First, more than 99% of completeness was primarily obtained in samples with Ct values ranging from 15.5 to 19.3, regardless of read count. The minimum number of reads to obtain >99% of completeness was 189, with a sample of Ct 15.5. The number of SARS-CoV-2 reads was confirmed to decrease with higher Ct values, with a negative correlation (*r* = -0.5, *p*-value = 0.008) between read counts and Ct values, yet read count alone was not sufficient to explain the decrease in completeness. As completeness also depends on average read length (**Figure 3**), we also observed a negative correlation (−0.47; *p*-value = 0.042) between Ct values and average read length for samples with Ct values >19. This result indicates that samples with higher Ct values may be unsuitable for such a method because of the fewer reads and the decreasing mean length of reads, precluding a full recovery of the genome sequences. Noticeably, samples associated with high viral loads (Ct<20) produced high completeness and GCS quality values (**Figure S2**; **Table S1**), even under multiplexing conditions (**Figure S3**). When multiplexing up to 12 samples on the same flow cell (**Figure S3**), even though up to 99% genome completeness could be reached for high viral load samples, base coverage was often <20, indicating insufficient sequencing depth for optimal GSC quality.

**Figure 3.**
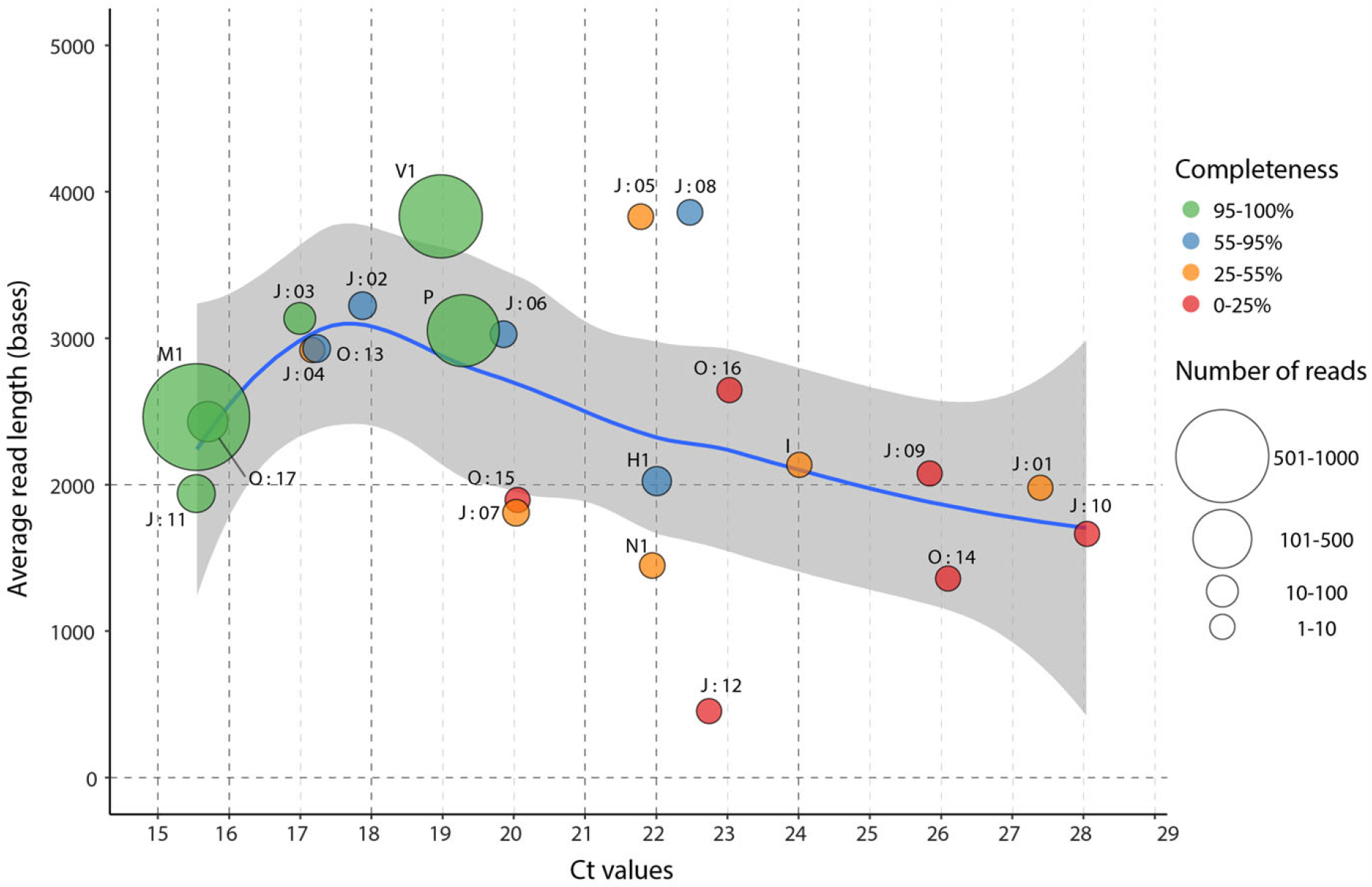
Relationships between average read length and Ct values. Each point represents a sequenced clinical sample. Samples that were multiplexed in a sequencing run are indicated by a colon followed by their barcode number. Symbol size is proportional to the obtained SARS-COV-2 read number and the color code indicates the proportions of the reference genome covered by at least one read (completeness). The blue line represents the fit of a local polynomial regression (loess) between average read lengths and Ct values for all samples in the study, and the grey area, its 95% confidence intervals.

### Validation of NASCarD in case of large genomic modifications in circulating genomes

Given that NASCarD is based on real-time mapping of translocating reads against a set of reference sequences (**Figure 1**), we investigated to which extent the choice of the reference sequences impacts the recovery of the target organism. We thus simulated six scenarios of divergent reference sequences by creating a range (i.e., 5, 10, 15, 20, 30, and 50%) of random single polymorphism nucleotides (SNP) *in silico* in the original omicron reference sequence (hCoV-19/USA/NY-NYS-34/2021; EPI_ISL ID 7887531) (**Figure 4**) (See Methods). We then mapped reads produced with NASCarD from one experiment (experiment V1; **Table S1**) presented previously (**Figure 2A**), as an exemplary case to this panel of modified reference sequences. All of the 870 reads generated in experiment V mapped to the omicron reference sequence the reference genome containing up to 5% SNPs. At 10% SNPs, we observed a small drop in the mapping rate to 98.9 %. We further mapped the reads to other SARS-CoV-2 references, including the original SARS-CoV-2 reference (Wuhan-Hu-1) and recombinant sub-lineage XE. In all cases, all reads could map to those alternative references (**Figure 4**). However, when reads were mapped onto the distantly related genome sequence of SARS coronavirus Tor2 (SARS-CoV-1; NC_004718.3), we observed that the mapping percentage dropped to 75.3%. Altogether, we thus provide evidence that the choice of genome references in the NASCarD approach for the case of SARS-CoV-2 can already cope with very large extents of diverging mutations and even recombinations, thus covering current and future, potential evolutionary events.

**Figure 4.**
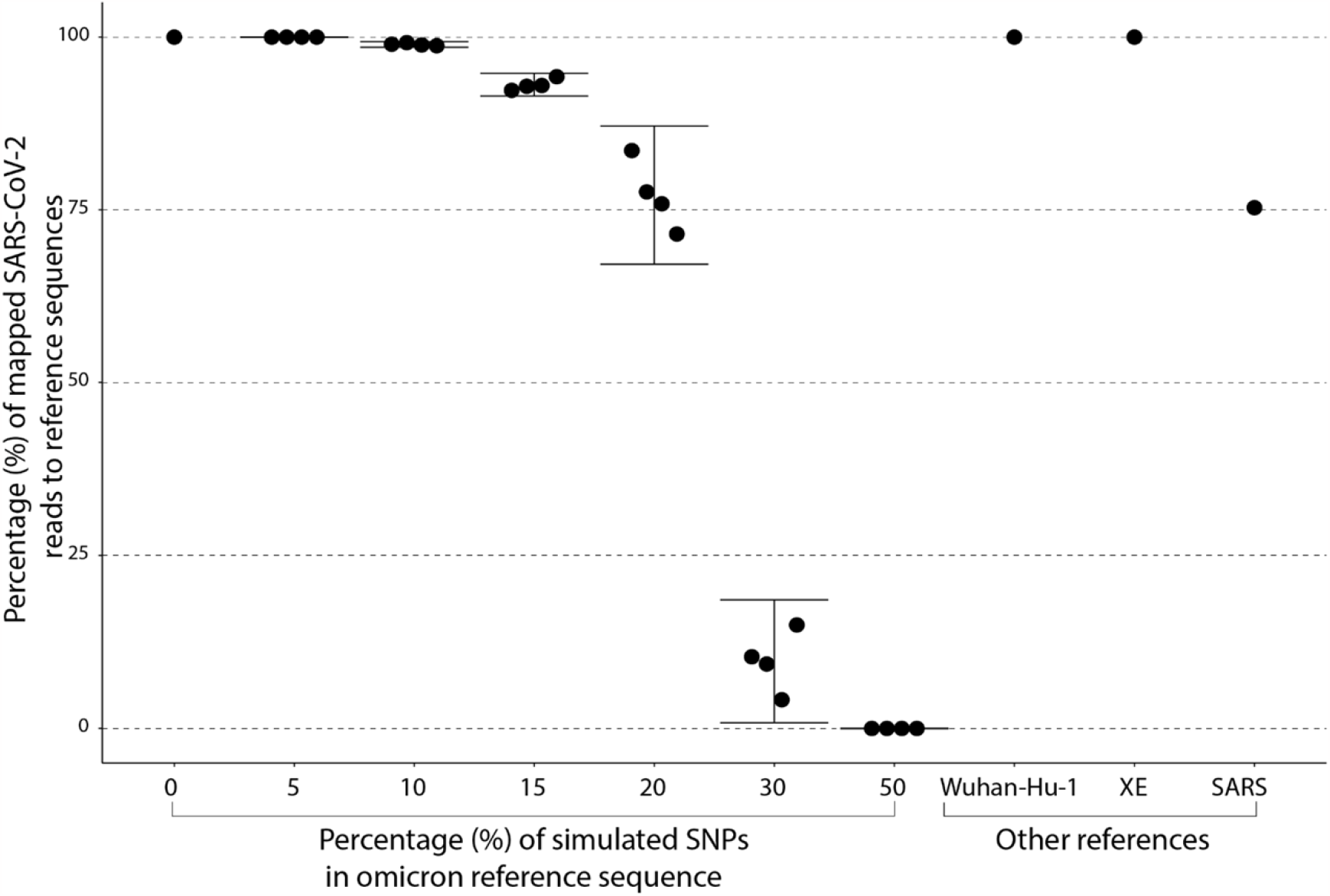
Percentage of experimental reads mapping to various genome reference sequences. *In silico* simulations were done to assess the capacity of the currently chosen set of SARS-CoV-2 reference sequences to capture novel mutations (Single Nucleotide Polymorphisms) and evolutionary events (recombinations) among circulating viruses. The vertical axis represents percentage of mapped reads from experiment V1 with respect to the chosen reference indicated on the horizontal axis. The 870 nanopore reads correspond to the totality of SARS-CoV-2 omicron reads identified in the sample, and not only to the ones enriched by AS. Additional references correspond to the sequences of isolate Wuhan-Hu-1 genome (NC_045512.2), a SARS-CoV-2 XE recombinant (EPI_ISL ID 13223243), and SARS-CoV-1 (NC_004718.3).

## DISCUSSION

The NASCarD approach allows low amounts of nucleic acids present in clinical samples to be specifically enriched during nanopore sequencing, limiting the costly and time-consuming production of non-target sequences in complex samples. The approach differs from standard nanopore sequencing approaches at the level of both library preparation (addition of carrier DNA) and sequencing (use of adaptive sampling) steps. In this study, we developed and validated NASCarD on clinical samples positive for SARS-CoV-2 during the Omicron wave, including Omicron recombinants, by investigating the completeness and quality of the generated consensus sequences, the turnaround time, and the cost associated with the new methodology. We showed that with NASCarD, high genome quality (>99% of bases with >20x coverage) and completeness could be obtained within seven hours of sequencing. The estimated costs of library preparation were also in the range of or cheaper than those of standard nanopore applications, especially because flow cells may be used for a shorter time with NASCarD as compared to standard nanopore applications (**Table S2**), and could potentially be washed and reused again.

To our knowledge, the combination of carrier DNA and AS to sequence directly from clinical samples, and specifically to target RNA viral pathogen, has not been done before. The use of a genomic carrier DNA was previously tested by adding bacterial (*Bacillus subtilis* ATCC 6633) DNA from a laboratory culture at low input concentrations to a background of Lambda carrier DNA (Mojarro et al. 2018), and further sequencing under standard nanopore sequencing conditions. Generally, unbiased shotgun metagenomics and metatranscriptomics are the most appropriate methods to identify novel emerging pathogens such as SARS-CoV-2 (Mokili, Rohwer, and Dutilh 2012). Our approach is unique among the latter, which have a longer turnaround time and associated costs (Charre et al. 2020; Quéromès et al. 2021; Bal et al. 2020). In addition, other studies have used nanopore AS in different contexts, with SARS-CoV-2, e.g., combined with PCR barcoding protocol from ONT (with 25 PCR cycles) to improve the recovery of SARS-CoV-2 viral genomes in ten clinical samples (Lin et al. 2022) or with customized sets of long amplicons (M. Wang et al. 2020), but also to enrich for specific bacterial species (Marquet et al. 2022; Martin et al. 2022; Gan et al. 2021; Payne et al. 2021; Bloomfield et al. 2023), but none have applied AS with successful SARS-CoV-2 whole genome sequencing without prior amplification.

The strengths of NASCarD are that the approach is simple, fast (because PCR-free) approach, flexible (run duration, number of samples), and is combined with real-time targeted genomic sequencing. It circumvents the need to have large amount of starting material for nanopore library preparation, an important technical limitation especially when considering the low amount of cDNA (<1 ng/*x03BC;l) typically retrieved from respiratory samples or other native clinical samples (Grädel et al. 2020; Lin et al. 2022). It also allows bypassing the need of designing PCR primers to produce overlapping amplicons across the viral genome. While the aforementioned approach allows for the generation of high-quality consensus sequences, it may not capture genetic diversity in regions that are difficult to amplify or yet unknown. This can result in missing or incomplete data for certain regions of the genome. Amplicon based approaches also require careful handling of PCR products to minimize contamination and ensure accurate sequencing results and interpretation. In terms of sensitivity, we demonstrated that, for high viral loads (Ct<20), the method may recover genome sequences of high quality, while allowing sample multiplexing to further reduce the cost per sample. Similarly, shotgun metagenomics has been successfully used on samples with Ct<20 (Quéromès et al. 2021; Bal et al. 2020), while for Ct in the range of 20-30 often found in clinical samples, the lower viral load or RNA quality of the samples (La Scola et al. 2020; Wölfel et al. 2020) may not be ideally suited for the NASCarD approach, as demonstrated here, if high genome completeness is required. Although SARS-CoV-2 detection in clinical samples is often reported in the range of 10-100 copies per ml using shotgun metagenomics (e.g. (Fauver et al. 2020; Sahajpal et al. 2021)), detection of few genomic fragments by those approaches may not provide sufficient in-depth genomic characterization of mutational variants in the circulating lineages. Besides shotgun metagenomics, other approaches such as SISPA (Chrzastek et al. 2022), which is based on an efficient method for non-targeted, random priming for whole genome sequencing of SARS-CoV-2, confirmed that limit of detections of 10^3^ pfu/ml (Ct 22) for whole genome assembly and 10^1^ pfu/ml (Ct 30) for metagenomic detection (Chrzastek et al. 2022) could be attained.

Some weaknesses of our study are that we analyzed samples from the delta and omicron waves in 2022 and did not consider the whole spectrum of SARS-CoV-2 lineages available. Yet, we demonstrated by *in silico* simulation that our reference sequences enable the future-proof detection of SARS-CoV-2 sequences even under very diverging evolutionary scenarios, as >98% of the nanopore reads from our clinical sample (omicron or recombinant lineages) could be retrieved even when using an AS reference sequence presenting up to 10% of SNP difference to the original sequence. Given its small genome size, availability in large quantities, low cost, and high purity, we choose the Lambda DNA as carrier DNA. Yet, this choice precludes applications where phage DNA or RNA are the main targets of the study. In that case, other sources of carrier DNA would need to be considered. Enriching for a small genome size such as that of SARS-CoV-2 is not problematic, but if the aim was to enrich for (or conversely deplete against) larger genomes, more latency in the AS decision could be foreseen. This latency is caused by the longer time needed for the initial 500 bases of a translocating read to be aligned to a large reference sequence, before deciding on reverting the voltage of the pore or not (Marquet et al. 2022; Ulrich et al. 2022),. This latency may then result in the sequencing of longer, non-targeted reads.

In conclusion, NASCarD significantly improves the targeted pathogen genomic sequencing directly from clinical samples and provides complete and accurate genome sequence reconstruction within seven hours at a competitive cost. We foresee that NASCarD may be combined with amplification-based WGS approach for SARS-CoV-2 lineage characterization in diagnostic laboratories. NASCarD can provide high-quality reference sequences for primer design, while surveillance of new variants may heavily benefit from amplicon-based genomic sequencing (e.g. (Charre et al. 2020)) or hybridization-based approaches (Nagy-Szakal et al. 2021), which may still provide sensitive sequencing at Ct>30. Future research should address how the new approach performs beyond SARS-CoV-2 sequencing, with other DNA or RNA pathogens present in clinical, surveillance and environmental samples.

## Data Availability

All data produced in the present work are contained in the manuscript and available online as indicated in the manuscript.

https://www.ncbi.nlm.nih.gov/bioproject/PRJNA930601

## SUPPLEMENTARY MATERIAL

**Figure S1**. Read length distribution between AS and control sequencing run for sample 1 experiment M using NASCarD.

**Figure S2**. Genome coverage of SARS-CoV-2 in COVID-19 positive clinical samples for experiments M, P, V, N, H, and I using NASCarD.

**Figure S3**. Genome coverage of 12 samples with different Ct values multiplexed in the same experiment using NASCarD.

**Table S1**. Summary of all experiments using NASCarD and control sequencing runs. Single-sample experiments are shaded in green; multiplex experiments are indicated in light blue.

**Table S2**. Sequencing costs and laboratory hands-on time.

## ETHICAL STATEMENT

The Ethics Committee of the Canton of Bern (BASEC-Nr 2022-00597) approved this study.

## ACKNOWLEDGEMENTS

The authors thank the diagnostic department of the Institute for Infectious Diseases, University of Bern for collecting and storing the samples.

