## Supplementary Material for "NASCarD (Nanopore Adaptive Sampling with Carrier DNA): A rapid, PCR-free method for whole genome sequencing of pathogens in clinical samples"

Figure S1. Read length distribution between AS and a control sequencing run for clinical sample 1 experiment M. A) After each run, all reads were consistently mapped to SARS-CoV-2, Lambda phage and human genomes. AS "Enriched" reads (red boxplot) mapped exclusively to SARS-CoV-2 yielding 1,197 reads with an average length of 2,462 bp. The "Rejected" reads correspond almost entirely to either human DNA (32,718 reads, in green) or Lambda DNA (639,328 reads, in blue) and have a mean length of 578 and 631, respectively. Moreover, only three reads with a size of 581 bp mapped to SARS-CoV-2. On the right side, control sequencing reads were mapped to SARS-CoV-2, yielding 745 reads with an average size of 2384 bp. In contrast to AS, the average size of both the human and Lambda genomes are 2796 bp and 8947 bp respectively. B) Comparison of mean maximum coverage for SARS-CoV-2 in AS (98.6 X) in red and control (59.4 X) in green after 52.4 hours of sequencing. C) SARS-CoV-2 genome completeness over time in AS and control sequencing as represented by 100 - %N. N represents position with a depth inferior to 20 X. In AS sequencing the genome completeness reaches more than 99% in 0.5 hours (red arrow) and in the control it reaches more than 99% in 0.3 hours (green arrow). The consensus quality in AS reaches 99.4 %, while in the control it only reaches a maximum of 97 %. Comparatively, the quality of the genome sequence using the standard PCR based approach was of 96.3% (i.e. 3.7% of Ns).

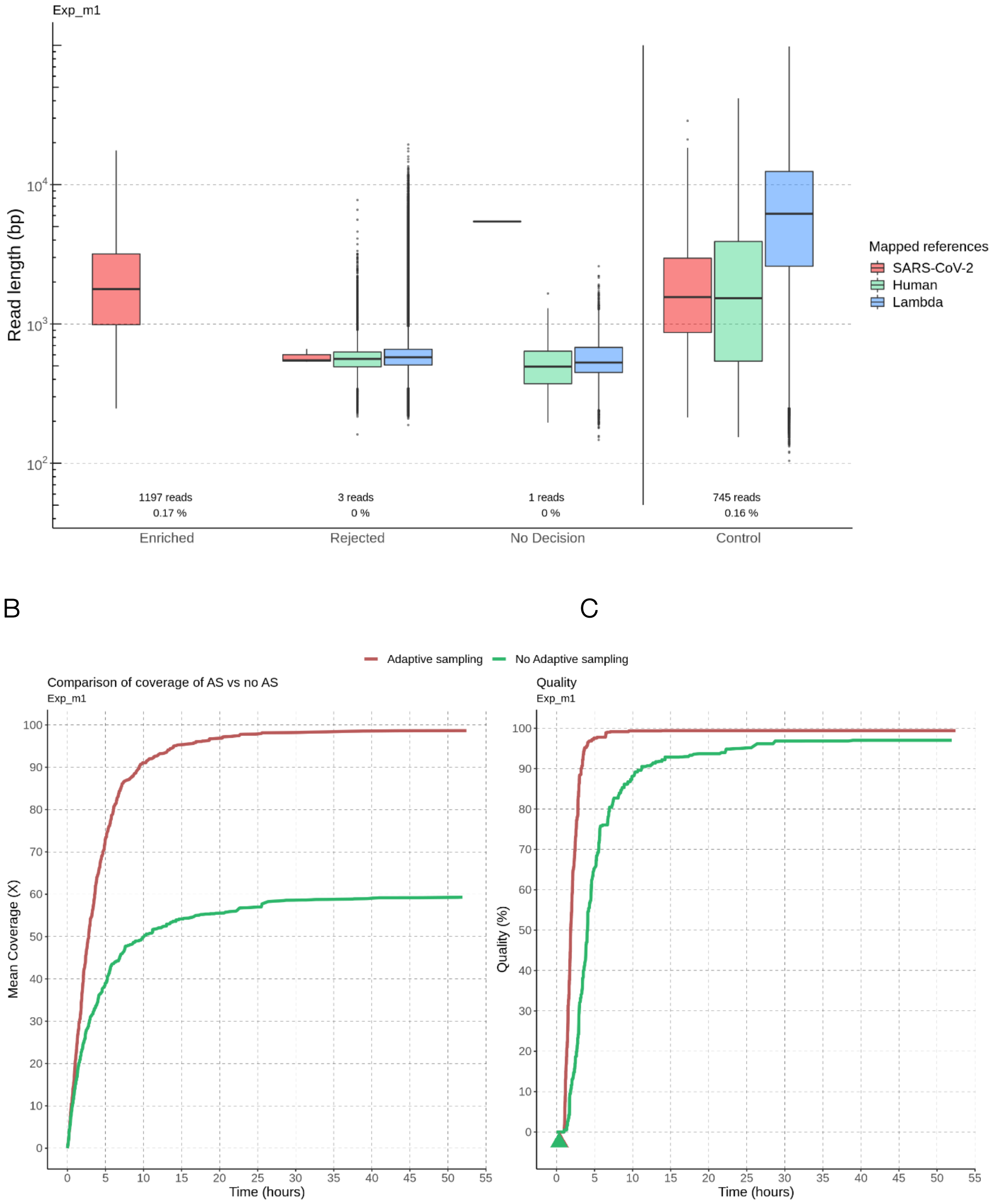

Figure S2. Genome coverage comparison of SARS-CoV-2 in COVID-19 positive clinical samples for experiments M, P, V, N, H, and I using NASCarD. Genome completeness (%) is indicated in each panel.

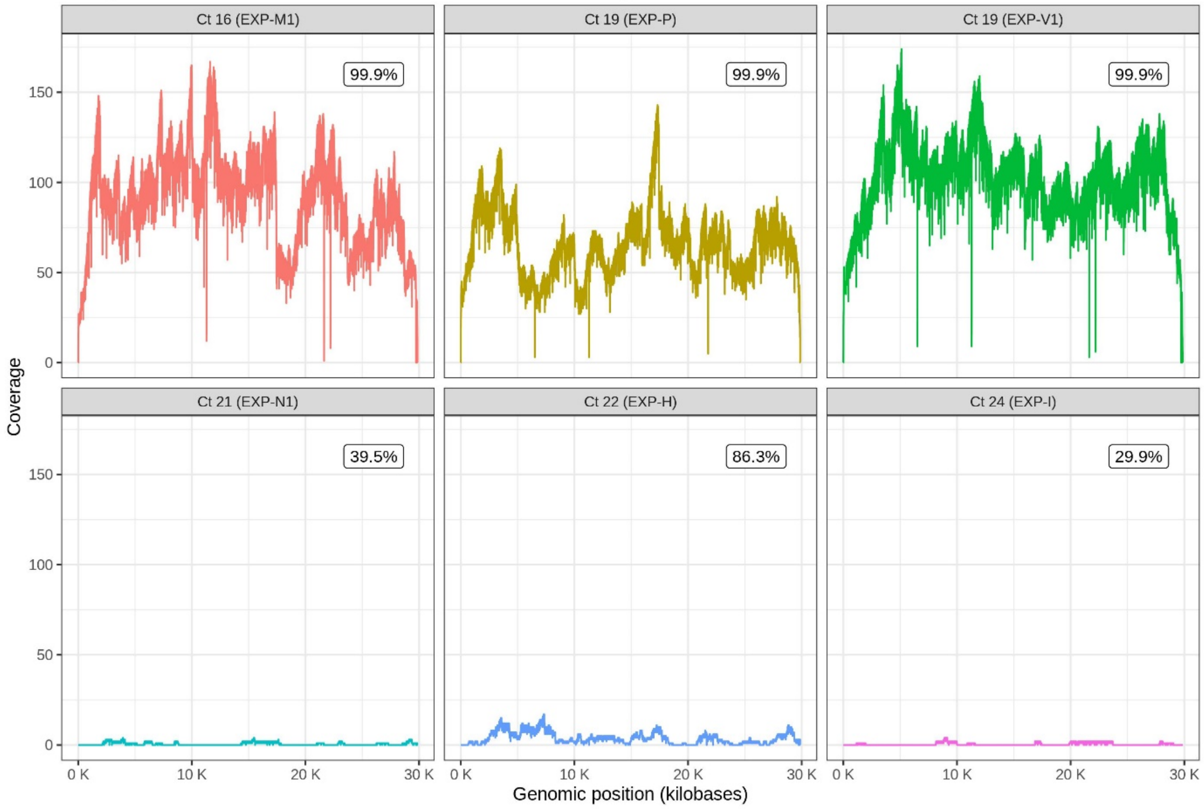

Figure S3. Genome coverage of 12 samples with different Ct values multiplexed in the same experiment using NASCarD. The sample with Ct 15.55 was also sequenced in a different individual experiment (experiment M1/M2; Figure S2). Genome completeness (%) is indicated in each panel.

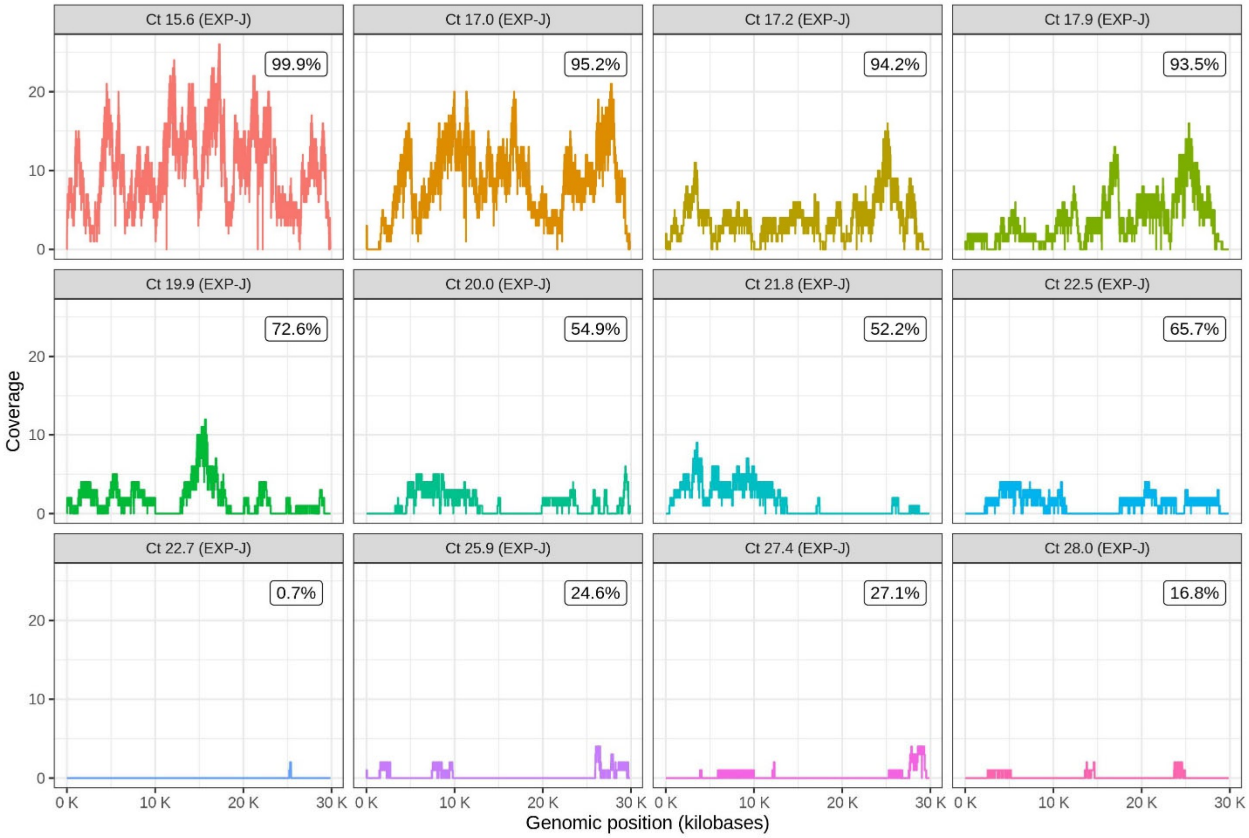

**Table S1.** Summary of all experiments using NASCarD and control sequencing runs. Single-sample experiments are shaded in green; multiplex experiments are indicated in light blue.

| | | | | | Sequencing results | | | | | | | | | | | |
|---|---|---|---|---|---|---|---|---|---|---|---|---|---|---|---|---|
| Experiment [a] | Sequencing method [b] | Sample identifier | Ct | Run duration (h) | Total | SARS-CoV-2 (% total) [c] | SARS-CoV-2 AS [d] | Human | Lambda | Unmapped | Mean read length ± standard deviation (bases) | Completeness (%) [e] | GCS quality | Mean coverage (x) | Lineage | GISAID ID |
| H | AS | H01 | 22.0 | 8 | 1,229,001 | 69 (0.0056%) | 65 | 372,308 | 853,063 | 3,561 | 2022.8 ± 1292.7 | 86.3 | 0 | 4.4 | BA.2 | EPI_ISL_13611368 |
| I | AS | I01 | 24.0 | 8 | 1,199,852 | 7 (0.0006%) | 7 | 213,976 | 966,079 | 19,790 | 2135.6 ± 2290.4 | 29.9 | 0 | 0.5 | BA.4 | EPI_ISL_13611359 |
| J | AS | J01 | 27.4 | 19 | 1,296,256 | 9 (0.0007%) | 8 | 12,700 | 113,714 | 212 | 1978.3 ± 1847.8 | 27.1 | 0 | 0.5 | BA.1 | EPI_ISL_10879502 |
| J | AS | J02 | 17.2 | 19 | 1,296,256 | 52 (0.004%) | 48 | 18,555 | 136,800 | 367 | 2929.2 ± 1875.4 | 94.2 | 0 | 4.7 | BA.2 | EPI_ISL_10879503 |
| J | AS | J03 | 17.9 | 19 | 1,296,256 | 44 (0.0034%) | 43 | 15,396 | 84,884 | 167 | 3222.7 ± 2163.8 | 93.5 | 0 | 4.6 | BA.2 | EPI_ISL_10879508 |
| J | AS | J04 | 17.0 | 19 | 1,296,256 | 99 (0.0076%) | 97 | 13,763 | 88,522 | 102 | 3136.3 ± 2455.7 | 95.2 | 0 | 10.2 | BA.2 | EPI_ISL_10879509 |
| J | AS | J05 | 21.8 | 19 | 1,296,256 | 16 (0.0012%) | 15 | 6,713 | 84,776 | 90 | 3830.3 ± 3634.5 | 52.2 | 0 | 1.9 | BA.1 | EPI_ISL_10879510 |
| J | AS | J06 | 19.9 | 19 | 1,296,256 | 29 (0.0022%) | 23 | 37,417 | 96,282 | 155 | 3027.2 ± 2578.2 | 72.6 | 0 | 2.3 | BA.1.1 | EPI_ISL_10879512 |
| J | AS | J07 | 20.0 | 19 | 1,296,256 | 26 (0.002%) | 25 | 52,737 | 74,338 | 157 | 1808.6 ± 2079.7 | 54.9 | 0 | 1.5 | BA.2 | EPI_ISL_10879518 |
| J | AS | J08 | 22.5 | 19 | 1,296,256 | 12 (0.0009%) | 12 | 6,382 | 93,880 | 106 | 3860.2 ± 2824.6 | 65.7 | 0 | 1.5 | BA.1.1 | EPI_ISL_10879520 |
| J | AS | J09 | 25.9 | 19 | 1,296,256 | 7 (0.0005%) | 7 | 14,008 | 81,315 | 183 | 2077.3 ± 1238.8 | 24.6 | 0 | 0.5 | BA.1 | EPI_ISL_10879522 |
| J | AS | J10 | 28.0 | 19 | 1,296,256 | 4 (0.0003%) | 4 | 9,758 | 72,601 | 88 | 1664.5 ± 950.1 | 16.8 | 0 | 0.2 | BA.2 | EPI_ISL_10879527 |
| J | AS | J11 | 15.6 | 19 | 1,296,256 | 207 (0.016%) | 189 | 12,725 | 102,352 | 138 | 1941.0 ± 1652.3 | 99.9 | 0 | 12.3 | BA.2 | EPI_ISL_10879533 |
| J | AS | J12 | 22.7 | 19 | 1,296,256 | 1 (0.0001%) | 1 | 12,038 | 52,218 | 111 | 456.0 ± 0 | 0.7 | 0 | 0 | BA.1.1 | EPI_ISL_10879538 |
| M1 | AS | M01 | 15.6 | 71.6 | 715,685 | 1,253 (0.1751%) | 1,197 | 39,437 | 674,483 | 512 | 2461.8 ± 2092.0 | 99.9 | 99.4 | 98.6 | BA.2 | EPI_ISL_10879533 |
| M2 | C | M01 | 15.6 | 72 | 454,012 | 745 (0.1641%) | 745 | 22,624 | 430,333 | 310 | 2383.6 ± 2583.1 | 99.9 | 97 | 59.4 | BA.2 | EPI_ISL_10879533 |
| N1 | AS | N01 | 21.9 | 71.8 | 485,087 | 14 (0.0029%) | 14 | 12,943 | 471,740 | 390 | 1448.1 ± 1216.9 | 39.5 | 0 | 0.6 | XM | EPI_ISL_12490028 |
| N2 | C | N01 | 21.9 | 71.6 | 320,728 | 10 (0.0031%) | 10 | 8,387 | 312,023 | 308 | 2386.5 ± 2082.8 | 37.3 | 0 | 0.8 | XM | EPI_ISL_12490028 |
| O | AS | O01 | 17.2 | 17.4 | 611,138 | 11 (0.0018%) | 10 | 25,896 | 131,804 | 422 | 2919.8 ± 2358.0 | 39.7 | 0 | 1 | BA.1.1 | EPI_ISL_11825556 |
| O | AS | O02 | 26.1 | 17.4 | 611,138 | 3 (0.0005%) | 2 | 754 | 102,161 | 71 | 1358.5 ± 726.2 | 5.0 | 0 | 0 | BA.1.1 | EPI_ISL_11825569 |
| O | AS | O03 | 20.1 | 17.4 | 611,138 | 5 (0.0008%) | 5 | 9,221 | 96,853 | 61 | 1895.4 ± 894.0 | 16.4 | 0 | 0.3 | BA.1.1 | EPI_ISL_11825577 |
| O | AS | O04 | 23.0 | 17.4 | 611,138 | 3 (0.0005%) | 3 | 1,640 | 101,600 | 49 | 2645.7 ± 2698.0 | 14.7 | 0 | 0.2 | BA.1.1 | EPI_ISL_11825599 |
| O | AS | O05 | 15.7 | 17.4 | 611,138 | 247 (0.0404%) | 235 | 24,863 | 115,243 | 231 | 2428.7 ± 1873.8 | 99.9 | 3.8 | 19.1 | AY.4 | EPI_ISL_6012191 |
| P1 | AS | P01 | 19.3 | 20.5 | 1,375,980 | 695 (0.0505%) | 693 | 74,532 | 1,300,753 | NA | 3051.5 ± 2408.4 | 99.9 | 99.7 | 70.8 | B.1.1.529 | EPI_ISL_7062525 |
| P2 | C | P01 | 19.3 | 20.5 | 432,020 | 179 (0.0414%) | 693 | 33,388 | 395,298 | 3155 | 3051.5 ± 2408.4 | 99.9 | 0.2 | 16 | B.1.1.529 | EPI_ISL_7062525 |
| V1 | AS | V01 | 19.0 | 18.5 | 2,989,153 | 870 (0.0291%) | 855 | 1,384,307 | 1,163,665 | 440,311 | 3833.6 ± 2757.3 | 99.9 | 99.7 | 109.7 | BA.2 | EPI_ISL_13251070 |
| V2 | C | V01 | 19.0 | 18.5 | 736,061 | 276 (0.0375%) | 276 | 337,429 | 320,678 | 77,678 | 3700.0 ± 5914.1 | 99.9 | 31.6 | 34.1 | BA.2 | EPI_ISL_13251070 |

[a] Experiments J and O consisted of multiplex runs, while all other experiments were performed as one sample per flow cell.
[b] "AS" refers to Adaptive sampling, while "C" refers to Control (without AS) sequencing method of the same library as prepared for AS.
[c] Total of all reads mapped to SARS-CoV-2 in the AS flow cell.
[d] Reads assigned to SARS-CoV-2 from the "stop receiving" data set in the AS flow cell.
[e] SARS-CoV-2 genome completeness is defined as the percentage of the corresponding genome sequence produced by amplicon-based sequencing.

Table S2. Sequencing costs and laboratory hands-on time.

| Sequencing method[a] | Cost per sample (Swiss Francs) | Library preparation time | Sequencing time[b] | Turnaround time |
|---|---|---|---|---|
| Standard sequencing | 1040 | 3 h | 10 h | 13 h |
| Standard sequencing (multiplexing 12 samples) | 251 | 3.5 h | >18 h | >22 h |
| NASCarD | 1040 | 3 h | 7 h | 10 h |
| NASCarD (multiplexing 12 samples) | 251 | 3.5 h | 7 h | 11 h |

[a]Cost estimates are based on Oxford Nanopore Technologies SQK-LSK109 library preparation and further sequencing on R9.4.1 flow cells. [b]Sequencing time to achieve more than 99% completeness per sample.
